# Adenoma detection rate in fecal immunochemical test positive colonoscopies: a population-based observational study

**DOI:** 10.1101/2020.08.31.20185124

**Authors:** Bernard Denis, Isabelle Gendre, Nicolas Tuzin, Anne Guignard, Philippe Perrin, Gabriel Rahmi

## Abstract

**Background and aims:** Neoplasia-related indicators, such as adenoma detection rate (ADR), are a priority in the quality improvement process for colonoscopy. Our aim was to assess and compare different detection and characterization indicators in fecal occult blood test (FOBT)-positive colonoscopies, to analyze the factors influencing their variance, and to propose benchmarks.

**Methods:** Retrospective analysis of prospectively collected data from all colonoscopies performed between 2007 and 2019 after a positive guaiac-based FOBT or a fecal immunochemical test (FIT) in the population-based colorectal cancer (CRC) screening program conducted in Alsace, part of the French national program. Detection indicators included ADR, NewADR (including proximal serrated lesions), mean number of adenomas per colonoscopy, and proximal serrated lesion detection rate. Characterization indicators included non-neoplastic polyp detection rate.

**Results:** Overall, 13.455 FIT-positive colonoscopies were performed by 116 endoscopists. The overall ADR was 57.6% (95%CI 56.8-58.5). For each 10 μg/g increase in fecal hemoglobin concentration, a 2% increase in ADR was observed. Endoscopists whose ADR was ≥55% were high detectors for all neoplasia, including proximal serrated lesions and number of adenomas. The non-neoplastic polyp detection rate was 39.5% in highest detectors (ADR >70%), significantly higher than in lower detectors (21.4%) (p<0.001). There was a strong correlation between detection and characterization indicators, e.g. between proximal serrated lesion and non-neoplastic polyp detection rates (Pearson = 0.73; p<0.01).

**Conclusions:** A single indicator, NewADR, including proximal serrated lesions, is enough to assess the neoplasia yield of colonoscopy provided the target standard is raised between 55% and 70% in FIT-positive colonoscopies (65-80% in men, 45-60% in women).

## INTRODUCTION

Most countries undertake colorectal cancer (CRC) screening programs with fecal occult blood test (FOBT), guaiac-based FOBT (gFOBT) or fecal immunochemical test (FIT), flexible sigmoidoscopy or colonoscopy, all effective in reducing CRC incidence and mortality.^1^ All these screening methods lead to colonoscopy allowing detection of early-stage CRCs and removal of neoplastic polyps (NPs). Colonoscopy is an operator-dependent examination: adenoma and polyp detection vary dramatically between endoscopists.^2,3^ High adenoma detection rate (ADR) and high polypectomy rate (PR) are associated with lower risk of post-colonoscopy CRC and fatal post-colonoscopy CRC.^4-7^ Measuring the neoplasia yield is therefore a priority in the quality improvement process for colonoscopy.^8,9^ ADR is the most commonly recommended neoplasia-related quality indicator.^8-10^ However, it is seldom measured in routine practice because it depends on the pathology report; it is prone to gaming and may prompt the endoscopist to take a “one and done” approach to polypectomy; finally, it ignores completely the serrated pathway, which might represent 10 to 25% of colorectal carcinogenesis.^11^ Other indicators have been proposed: mean number of adenomas (MNA) per colonoscopy, which is a better reflect of full length of colon examination;^3,8^ mean number of adenomas per positive colonoscopy (MNAPPC), which is correlated with adenoma and advanced adenoma miss rates;^12^ polyp detection rate (PDR), equivalent to PR, which is easy to calculate as it can be derived from administrative data;^13^ and proximal serrated lesion detection rate (ProxSLDR), which assesses the serrated pathway.^11^

Despite many published studies about ADR, there is still no standardized way to calculate it. The latest US recommendations are the first to specify that sessile serrated lesions (SSLs) “should not be counted toward the ADR”.^8^ ADR calculation method varies greatly across studies and is often not specified, even in landmark studies.^4,6,7^ Depending on studies, CRCs and SSLs are counted, or not, and incomplete colonoscopies are included, or not, for the calculation of ADR. These different calculation methods hamper comparison between series and screening programs.^14^ There is a large body of literature about ADR in the setting of screening colonoscopies, but literature is scarce in the setting of FOBT-positive colonoscopies, particularly for FIT.^15-19^ The US Multi-Society Task Force on CRC recommended that ADR should be greater than 45% in men and 35% in women in FIT-positive colonoscopies with a hemoglobin threshold of 20 *μ*g/g or less (weak recommendation; very low quality of evidence).^20^ The European Society of Gastrointestinal Endoscopy (ESGE) recommendations stipulated that “in FIT-positive enriched populations, the minimum standard may need to be higher than 25%; however, the exact value is yet to be established”.^9^

The benefit of the removal of NPs is diminished by the removal of non-neoplastic polyps (NNPs), mainly hyperplastic polyps (HPs) that account for around 22.5%-30% of polyps (75% of serrated polyps that account for 30%-40% of polyps).^21^ Accurate real-time endoscopic characterization of the histology of colorectal polyps would be crucial to determine whether a polyp has to be removed and analyzed or not. Optical technologies, such as narrow-band imaging (NBI), have been developed to help endoscopists differentiate adenomatous from non-adenomatous polyps, so that small non-adenomatous polyps can be safely left *in situ* without polypectomy. Their performances have been thoroughly investigated, mainly in expert hands, and are diversely appreciated.^22^ Performance levels for community endoscopists have been disappointing.^23^

The aim of this study was (1) to evaluate and compare different neoplasia-related detection and characterization indicators for colonoscopy and their calculation method, (2) to compare their results between gFOBT- and FIT-positive colonoscopies, (3) to analyze the factors influencing their variance, and (4) to establish minimum and target standards for the French CRC screening program with FIT.

## PATIENTS AND METHODS

### Screening program

The French organized CRC screening program, implemented from 2003, moved from gFOBT (Hemoccult II) to FIT (OC-Sensor) in 2015. Its design has been previously described.^24^ Residents aged 50-74 years are invited by mail every other year to participate. People with serious illness, recent CRC screening or high CRC risk are excluded. The FIT positivity threshold is set at 30 μg hemoglobin per gram (μg/g) feces so that the positivity rate would be 4 to 5%. People with a positive FOBT are referred for colonoscopy.

### Colonoscopies

All data concerning gFOBT-positive colonoscopies performed between 2007 and 2014 and FIT-positive colonoscopies performed between 2015 and 2019 within the screening program in Alsace, a region in eastern France (0.57 million residents aged 50-74), were prospectively collected and retrospectively analyzed. All certified endoscopists participated in the program. Colonoscopies were performed by community gastroenterologists, generally with sedation/anesthesia provided by an anesthesiologist. All colonoscopies were included, either complete to the cecum or not, whatever the quality of bowel preparation. Endoscopists who had performed <30 FOBT-positive colonoscopies were excluded for the assessment of neoplasia-related indicators.

### Pathological classification

The pathological examination of detected polyps was performed as routine procedure by community general pathologists. The result of each colonoscopy was classified according to the worst prognosis lesion. Conventional adenoma was defined as any tubulous, tubulo-villous or villous adenoma. Advanced adenoma (AA) was defined as a conventional adenoma measuring ≥10 mm or with a villous component >20% or with high-grade dysplasia. Serrated lesions (SLs) included SSLs, traditional serrated adenomas and HPs. NP was defined as any pre-cancerous lesion, including conventional adenoma, SSL and traditional serrated adenoma. NNP was defined as any non-precancerous lesion, including HP, normal or inflammatory mucosa, lipoma, lymph node, etc. Non-adenomatous non-serrated lesions (NANSL) included all normal or inflammatory mucosa, lipoma, lymph node, etc.

### Indicators

Colonoscopies displaying invasive CRC were excluded for the calculation of all neoplasia-related indicators. ADR, A10+DR, NPDR, AADR, SSLDR, PDR, NNPDR and NANSLDR were defined respectively as the percentages of colonoscopies where at least one conventional adenoma, one conventional adenoma ≥10 mm, one NP, one AA, one SSL, one polyp, one NNP and one NANSL were found. All SLs were excluded when calculating ADR, AADR and MNA.^8^ The distal HP detection rate (DistHPDR) was defined as the percentage of colonoscopies where at least one HP was found in the rectum or distal colon (distally to the splenic flexure, splenic flexure being included). The ProxSLDR was defined as the percentage of colonoscopies where at least one SL of any size was found in the proximal colon (proximally to the splenic flexure, splenic flexure being excluded).^25^ The NewADR was defined as the percentage of colonoscopies where at least one conventional adenoma was found or one SL of any size in the proximal colon. MNA, MNP and MNNP were defined respectively as the overall number of conventional adenomas, polyps, and NPs detected divided by the number of colonoscopies performed. The MNAPPC was defined as the number of conventional adenomas detected divided by the number of colonoscopies where at least one adenoma was detected. Neoplasia-related indicators were classified in two categories (table 3): detection (ADR, A10+DR, NewADR, NPDR, AADR, PDR, MNA, MNP, MNAPPC, SSLDR, ProxSLDR) and characterization indicators (NNPDR, DistHPDR, NANSLDR).

### Statistical methods

Qualitative variables were described by their frequency with 95% confidence intervals (95% CI) and quantitative variables by their median, range, mean and standard deviation (SD). The relationship between neoplasia-related quality indicators was evaluated using the Pearson correlation coefficient *r*. The chi-square test was used to search for statistical significance by comparisons of proportions. Multivariate analyses (binomial mixed regressions) were performed to explore the factors influencing ADR, NewADR, A10+DR, ProxSLDR, and NNPDR. The following factors were analyzed: sex, age, year, screening history, screening test (gFOBT vs FIT), fecal hemoglobin concentration, time to colonoscopy, endoscopist, private or hospital practice, annual volume of FOBT-positive colonoscopies, and cecal intubation rate (CIR). The significance level was set at 0.05. Statistical analyses were performed using R software version 3.6.0.

All authors had access to the study data and reviewed and approved the final manuscript. This study was exempt from institutional review board approval because individual participants were not approached, only routinely collected data were utilised and all data were anonymized for the purposes of quality improvement within the screening program. The study was performed in accordance with the declaration of Helsinki.

## RESULTS

The FIT uptake was 44.4% in 2018-19 and the positivity rate was 3.8%. During the study period, 14,228 individuals had FIT-positive colonoscopies performed by 116 endoscopists. Among these individuals, 773 (5.4%) had colonoscopies displaying a CRC and were excluded, so that the colonoscopies of 13,455 individuals (mean age 62.4 years (standard deviation (SD) 7.0); men 59.6%) were analyzed. During these procedures 23,379 polyps were removed, the characteristics of which are detailed in supplementary file 1. The overall CIR was 97.8% (mean 96.1%; SD 4.8%): ≥90% for 94.0% of endoscopists and ≥95% for 80.2%.

### Colonoscopy volume

The number of colonoscopies per endoscopist varied from 1 to 623 (mean 116, SD 126, median 85). The median time-frame for reaching 30 FIT-positive colonoscopies was 9 months; it was 23 months for reaching 100. The CIR was ≥90% for 87.5% of low-volume endoscopists (<20 annual FIT-positive colonoscopies), significantly lower than the 98.5% recorded among high-volume endoscopists (p=0.01). There was no correlation between endoscopist’s colonoscopy volume and all neoplasia-related indicators except for ProxSLDR and NANSLDR. ProxSLDR was 5.9% in 42 (36.2%) low-volume endoscopists, significantly lower than 7.8% in high-volume endoscopists (p=0.01). NANSLDR was 8.8% in low-volume endoscopists, significantly higher than 6.2% in high-volume endoscopists (p=0.0006).

A total of 388 colonoscopies performed by 36 endoscopists were excluded for further analysis as these practitioners had done <30 colonoscopies during the study period. Consequently, 13,067 colonoscopies were finally evaluated, performed by 80 endoscopists. Table 1 shows the results of detection and characterization indicators and their benchmark rates recommended in France. Table 2 shows the correlations between the values of detection and characterization indicators.

**Table 1:**
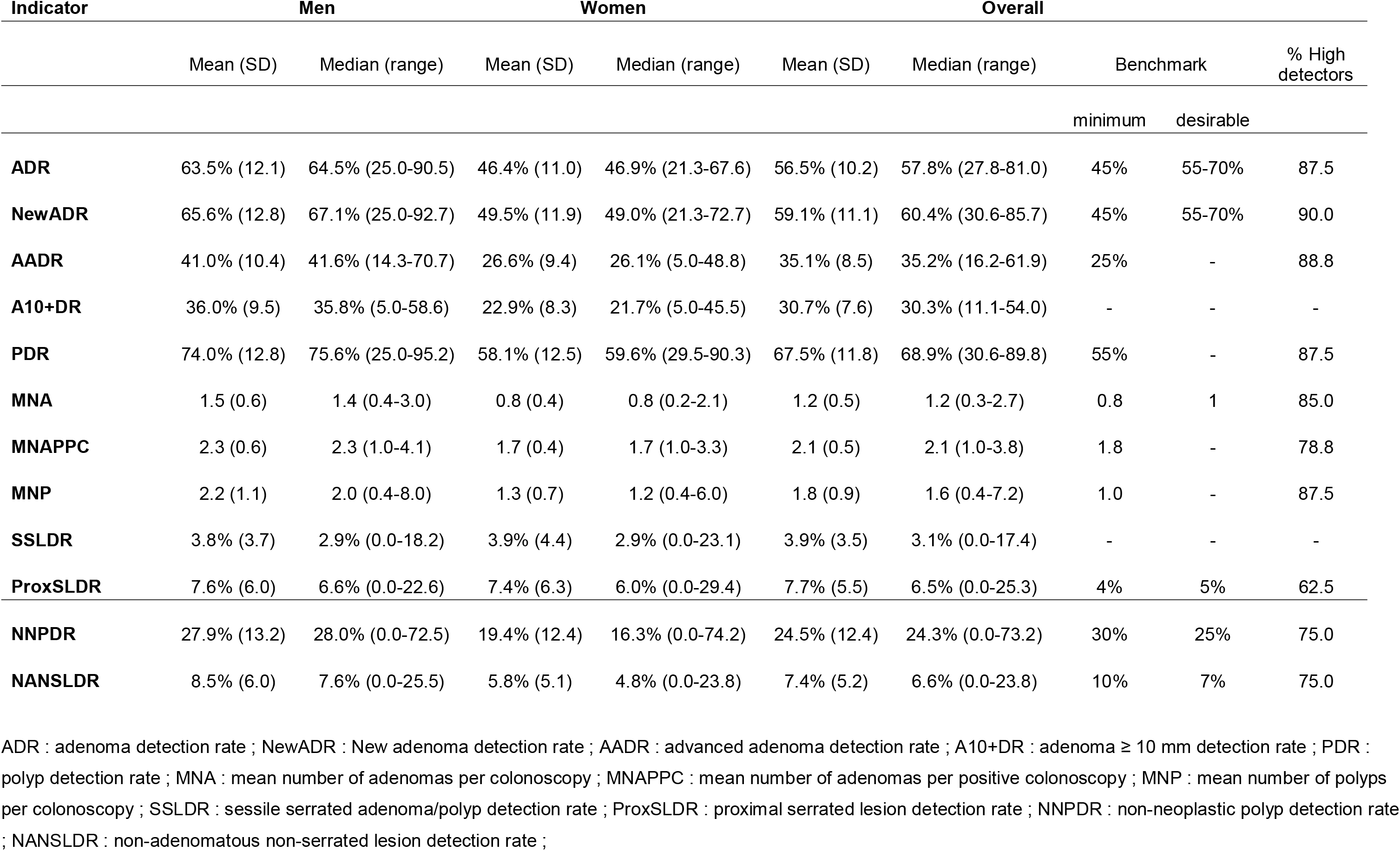
Detection and characterization indicators within the fecal immunochemical test screening program (80 endoscopists having performed ≥ 30 colonoscopies)

| Indicator | Men |  | Women |  | Overall |  |  |  |  |
| --- | --- | --- | --- | --- | --- | --- | --- | --- | --- |
|  | Mean (SD) | Median (range) | Mean (SD) | Median (range) | Mean (SD) | Median (range) | Benchmark |  | % High detectors |
|  |  |  |  |  |  |  | minimum | desirable |  |
| ADR | 63.5% (12.1) | 64.5% (25.0-90.5) | 46.4% (11.0) | 46.9% (21.3-67.6) | 56.5% (10.2) | 57.8% (27.8-81.0) | 45% | 55-70% | 87.5 |
| NewADR | 65.6% (12.8) | 67.1% (25.0-92.7) | 49.5% (11.9) | 49.0% (21.3-72.7) | 59.1% (11.1) | 60.4% (30.6-85.7) | 45% | 55-70% | 90.0 |
| AADR | 41.0% (10.4) | 41.6% (14.3-70.7) | 26.6% (9.4) | 26.1% (5.0-48.8) | 35.1% (8.5) | 35.2% (16.2-61.9) | 25% | - | 88.8 |
| A10+DR | 36.0% (9.5) | 35.8% (5.0-58.6) | 22.9% (8.3) | 21.7% (5.0-45.5) | 30.7% (7.6) | 30.3% (11.1-54.0) | - | - | - |
| PDR | 74.0% (12.8) | 75.6% (25.0-95.2) | 58.1% (12.5) | 59.6% (29.5-90.3) | 67.5% (11.8) | 68.9% (30.6-89.8) | 55% | - | 87.5 |
| MNA | 1.5 (0.6) | 1.4 (0.4-3.0) | 0.8 (0.4) | 0.8 (0.2-2.1) | 1.2 (0.5) | 1.2 (0.3-2.7) | 0.8 | 1 | 85.0 |
| MNAPPC | 2.3 (0.6) | 2.3 (1.0-4.1) | 1.7 (0.4) | 1.7 (1.0-3.3) | 2.1 (0.5) | 2.1 (1.0-3.8) | 1.8 | - | 78.8 |
| MNP | 2.2 (1.1) | 2.0 (0.4-8.0) | 1.3 (0.7) | 1.2 (0.4-6.0) | 1.8 (0.9) | 1.6 (0.4-7.2) | 1.0 | - | 87.5 |
| SSLDR | 3.8% (3.7) | 2.9% (0.0-18.2) | 3.9% (4.4) | 2.9% (0.0-23.1) | 3.9% (3.5) | 3.1% (0.0-17.4) | - | - | - |
| ProxSLDR | 7.6% (6.0) | 6.6% (0.0-22.6) | 7.4% (6.3) | 6.0% (0.0-29.4) | 7.7% (5.5) | 6.5% (0.0-25.3) | 4% | 5% | 62.5 |
| NNPDR | 27.9% (13.2) | 28.0% (0.0-72.5) | 19.4% (12.4) | 16.3% (0.0-74.2) | 24.5% (12.4) | 24.3% (0.0-73.2) | 30% | 25% | 75.0 |
| NANSLDR | 8.5% (6.0) | 7.6% (0.0-25.5) | 5.8% (5.1) | 4.8% (0.0-23.8) | 7.4% (5.2) | 6.6% (0.0-23.8) | 10% | 7% | 75.0 |

ADR : adenoma detection rate ; NewADR : New adenoma detection rate ; AADR : advanced adenoma detection rate ; A10+DR : adenoma $\geq 10$ mm detection rate ; PDR : polyp detection rate ; MNA : mean number of adenomas per colonoscopy ; MNAPPC : mean number of adenomas per positive colonoscopy ; MNP : mean number of polyps per colonoscopy ; SSLDR : sessile serrated adenoma/polyp detection rate ; ProxSLDR : proximal serrated lesion detection rate ; NNPDR : non-neoplastic polyp detection rate ; NANSLDR : non-adenomatous non-serrated lesion detection rate ;

**Table 2:** Correlations between the values of detection and characterization indicators in the fecal immunochemical test screening program (80 endoscopists having performed ≥ 30 colonoscopies)

|  |  | Pearson (p) – R2 |  |  |  |  |  |  |  |  |
| --- | --- | --- | --- | --- | --- | --- | --- | --- | --- | --- |
| **ADR** | 1.00 - 1.00 |  |  |  |  | **NNPDR** | 1.00 - 1.00 |  |  |  |
| **NewADR** | **0.99 (p<0.01) - 0.98** | 1.00 - 1.00 |  |  |  | **DistHPDR** | **0.95 (p<0.01) - 0.91** | 1.00 - 1.00 |  |  |
| **AADR** | 0.76 (p<0.01) - 0.57 | 0.76 (p<0.01) - 0.58 | 1.00 - 1.00 |  |  | **NANSLDR** | 0.72 (p<0.01) - 0.52 | 0.53 (p<0.01) - 0.28 | 1.00 - 1.00 |  |
| **A10+DR** | **0.79 (p<0.01) - 0.62** | **0.80 (p<0.01) - 0.64** | 0.86 (p<0.01) - 0.74 | 1.00 - 1.00 |  |  | **NNPDR** | **DistHPDR** | **NANSLDR** | 1.00 - 1.00 |
| **PDR** | **0.94 (p<0.01) - 0.88** | **0.95 (p<0.01) - 0.91** | 0.71 (p<0.01) - 0.51 | 0.79 (p<0.01) - 0.63 | 1.00 - 1.00 |  |  |  |  |  |
| **MNA** | **0.87 (p<0.01) - 0.76** | **0.87 (p<0.01) - 0.76** | 0.61 (p<0.01) - 0.38 | **0.68 (p<0.01) - 0.47** | **0.89 (p<0.01) - 0.79** | 1.00 - 1.00 |  |  |  |  |
| **MNAPPC** | **0.69 (p<0.01) - 0.48** | **0.69 (p<0.01) - 0.48** | 0.46 (p<0.01) - 0.21 | **0.56 (p<0.01) - 0.31** | **0.75 (p<0.01) - 0.57** | 0.94 (p<0.01) - 0.88 | 1.00 - 1.00 |  |  |  |
| **MNP** | 0.72 (p<0.01) - 0.52 | 0.73 (p<0.01) - 0.53 | 0.51 (p<0.01) - 0.26 | **0.61 (p<0.01) - 0.37** | 0.79 (p<0.01) - 0.62 | **0.90 (p<0.01) - 0.81** | 0.88 (p<0.01) - 0.78 | 1.00 - 1.00 |  |  |
| **SSLDR** | 0.53 (p<0.01) - 0.28 | 0.57 (p<0.01) - 0.32 | 0.39 (p<0.01) - 0.15 | **0.38 (p<0.01) - 0.14** | 0.55 (p<0.01) - 0.30 | 0.62 (p<0.01) - 0.38 | 0.49 (p<0.01) - 0.24 | 0.44 (p<0.01) - 0.19 | 1.00 - 1.00 |  |
| **ProxSLDR** | 0.73 (p<0.01) - 0.53 | 0.78 (p<0.01) - 0.61 | 0.49 (p<0.01) - 0.24 | **0.57 (p<0.01) - 0.33** | 0.77 (p<0.01) - 0.60 | **0.80 (p<0.01) - 0.63** | 0.67 (p<0.01) - 0.45 | 0.73 (p<0.01) - 0.54 | 0.77 (p<0.01) - 0.59 | 1.00 - 1.00 |
|  | **ADR** | **NewADR** | **AADR** | **A10+DR** | **PDR** | **MNA** | **MNAPPC** | **MNP** | **SSLDR** | **ProxSLDR** |

### Detection indicators

During the FIT period, the overall ADR was 57.6% (95%CI 56.8-58.5), 58.0% in the first round and 57.0% in the second (p=0.2). Individual endoscopists’ ADR ranged from 27.8% to 81.0% (mean 56.5%, SD 10.2%). Of 72 endoscopists having an ADR ≥45%, 5 (6.9%) had an MNA below 0.8, whereas of 48 endoscopists having an ADR ≥55%, only one (2.1%) had an MNA below 0.8 (fig 1). Likewise, for MNAPPC (13.9% vs 2.1%, respectively; cut-off 1.8) and for ProxSLDR (36.1% vs 4.2%, respectively; cut-off 4%) (fig 2). Overall, 40.0% of the endoscopists had an ADR ≤55% and 52.5% an ADR between 55% and 70%. Overall, 4828/13455 (35.9%) patients had their colonoscopies performed by 56/116 (48.3%) endoscopists who either had a CIR <90% or an ADR <55% (1072 (8.0%) patients by 26/116 (22.4%) endoscopists having CIR <90% or ADR <45%).

**Figure 1:**
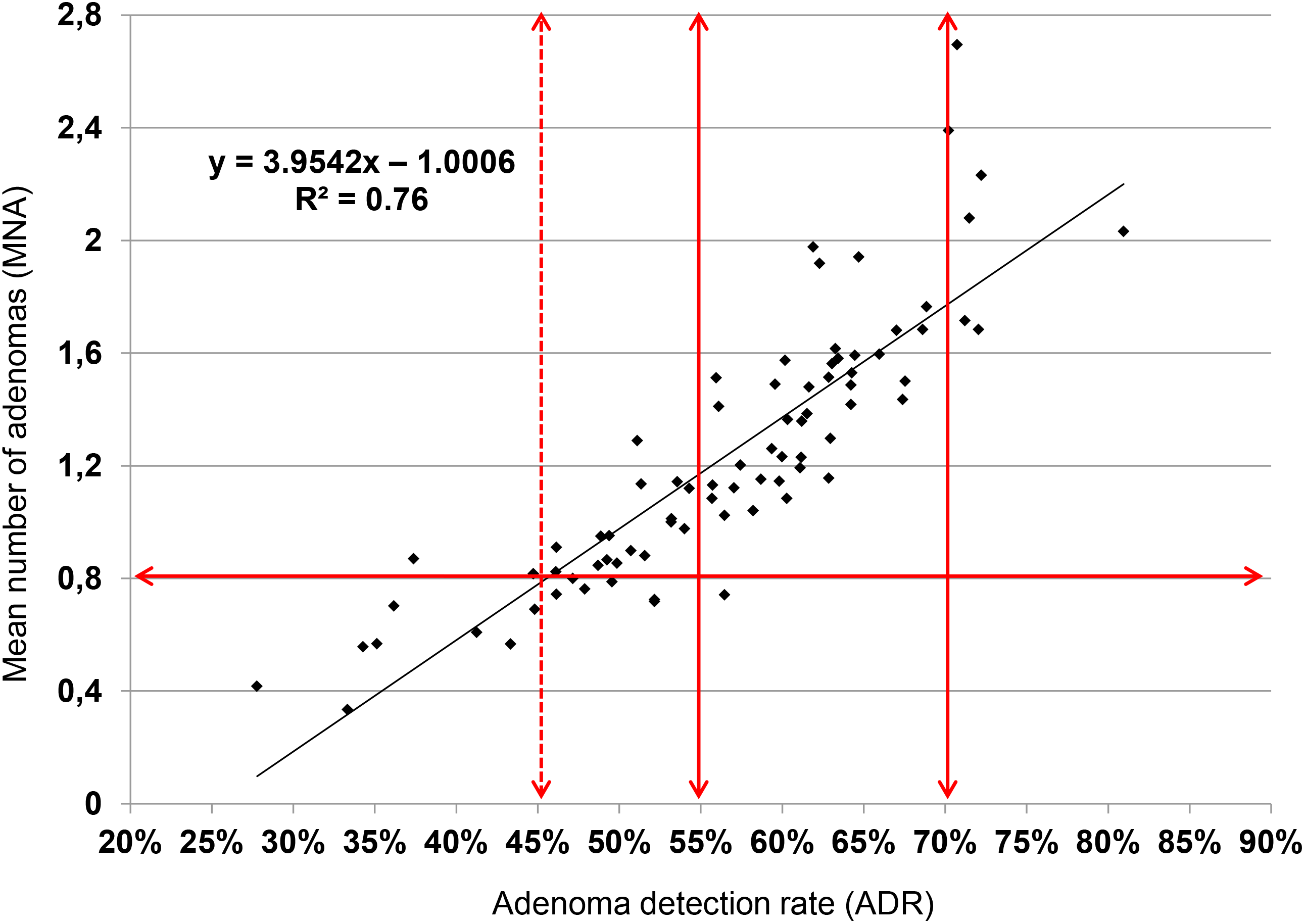
Correlation between adenoma detection rate and mean number of adenomas per colonoscopy

**Figure 2:**
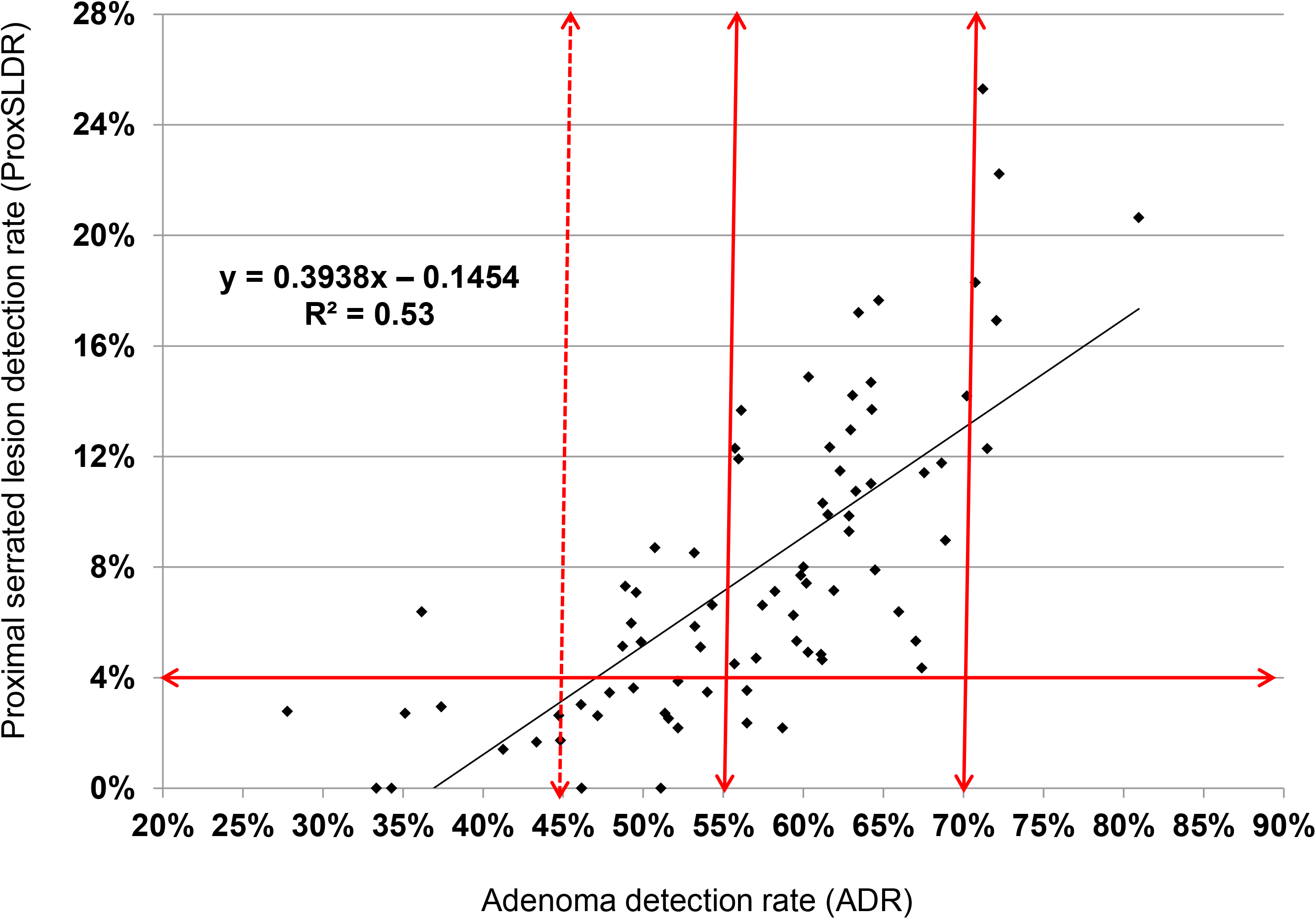
Correlation between adenoma and proximal serrated lesions detection rates

The way ADR was calculated modified the results. The overall NewADR, including proximal SLs, was 60.2%, significantly higher than the overall ADR that excluded them (57.6%, p≤0.001) (difference from 0 to 8.7% depending on the endoscopist). Depending on the ADR cut-off adopted, 2.5 to 7.5% of endoscopists would become high detectors if proximal SLs were included. If the overall ADR were calculated including CRCs, it would have been 59.9%, significantly higher than the overall ADR calculated excluding them (57.6%, p<0.001) (difference from 0 to 4.9% depending on the endoscopist). Depending on the ADR cut-off, 2.5 to 5% of endoscopists would become high detectors if CRCs were included. By contrast, the overall ADR calculated on colonoscopies complete to the cecum only was 58.0%, not significantly different from the ADR calculated on all colonoscopies (57.6%, p=0.6) (difference between 0 and 4.9% depending on the endoscopist).

Globally, 0.84 × PDR gave an estimate of ADR. However, individually, the ADR/PDR ratio varied from 0.61 to 0.93 depending on the endoscopist (supplementary file 2).

### Characterization indicators

Most NNPs were distal HPs (mean 16.2%) and NANSLs (mean 7.4%). Overall, the positive predictive value (PPV) of optical diagnosis for NPs was 72.8% (MNNP/MNP), varying from 37.8% to 100% depending on the endoscopist. It was inversely significantly correlated with all other characterization indicators: e.g. DistHPDR (*r* = – 0.69; p≤0.01; coefficient of determination R2=0.47) and NNPDR (*r* = – 0.68; p≤0.01; R2=0.46). The NNPDR was 39.5% in endoscopists whose ADR was >70%, significantly higher than 21.4% in those whose ADR was ≤70% (p<0.001). The correlation between characterization (NNPDR) and detection indicators was moderate for ADR (*r* = 0.64; p<0.01; R2=0.40) (Fig 3) and strong for ProxSLDR (*r* = 0.73; p<0.01; R2=0.54).

**Figure 3:**
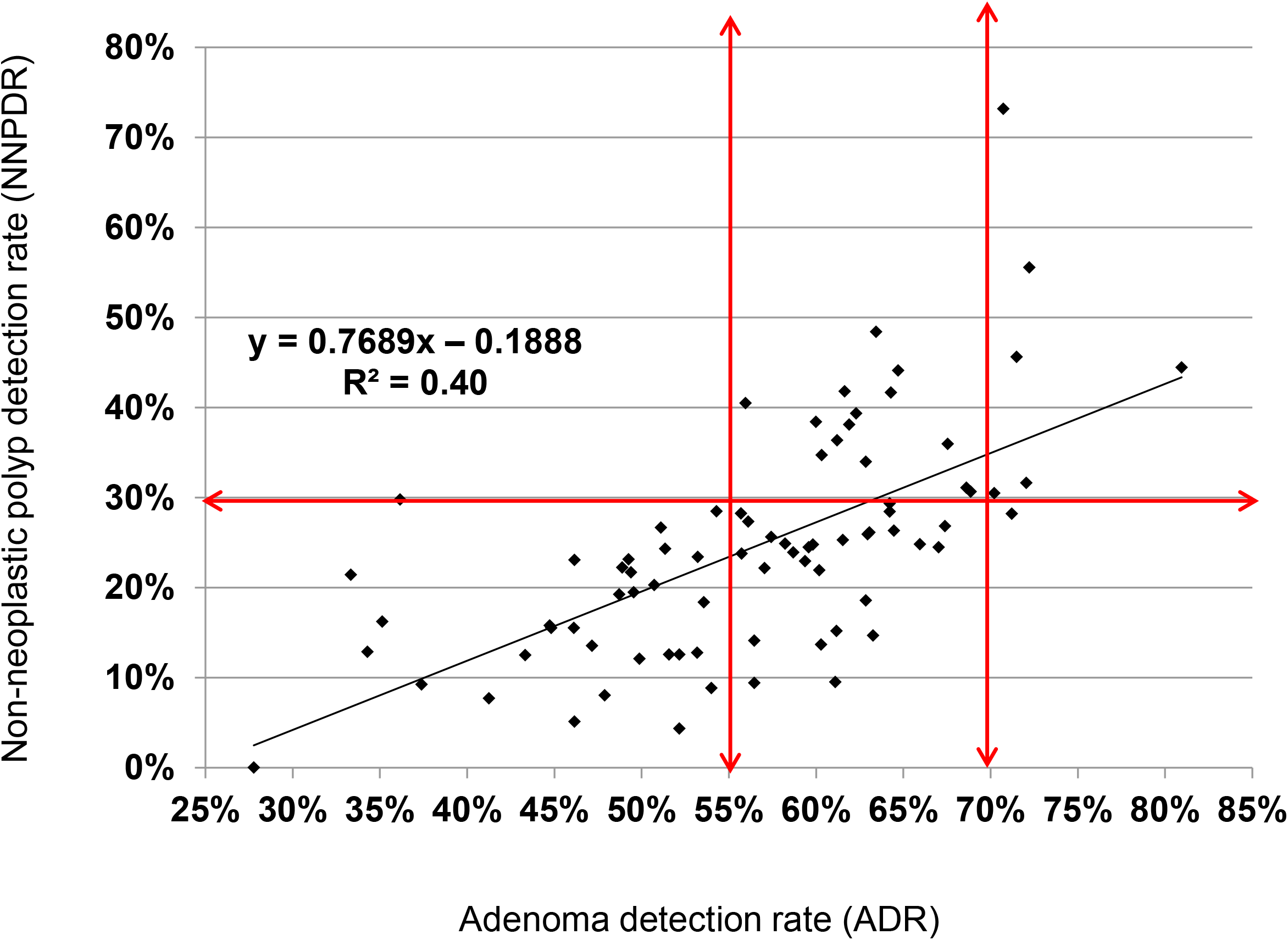
Correlation between adenoma and non-neoplastic polyps detection rates

### Time course of detection and characterization indicators

Conventional adenomas detection indicators increased significantly between gFOBT and FIT periods but remained stable within each of the two periods (supplementary files 3 and 4). By contrast, ProxSLDR increased significantly and continuously throughout the whole study (e.g. from 7.0% to 8.5% between the two FIT rounds (p=0.002)). Likewise, characterization indicators increased significantly throughout the whole study (e.g. NNPDR from 15.4% to 22.9% (p<0.001)).

### Factors influencing detection and characterization indicators (table 3 and supplementary file 5)

In multivariate analysis, the factors influencing ADR, NewADR, and A10+DR significantly were sex, age, screening history, type of FOBT, fecal hemoglobin concentration, CIR <90%, and endoscopist. A 10 μg/g increase in fecal hemoglobin concentration was associated with a 2% increase in ADR. Private or public practice, year of colonoscopy, time to colonoscopy, and FIT-positive colonoscopy volume were not significantly associated (data not shown). The factors influencing ProxSLDR significantly were previous colonoscopy screening, type of FOBT, CIR <90% and endoscopist. Those influencing NNPDR were sex, previous colonoscopy screening, type of FOBT and endoscopist.

**Table 3:** Multivariate analyses of factors influencing detection and characterization indicators during the fecal immunochemical test screening period

|  | **ADR** |  | **NewADR** |  | **A10+DR** |  | **ProxSLDR** |  | **NNPDR** |  |
| --- | --- | --- | --- | --- | --- | --- | --- | --- | --- | --- |
| Determinants | Odds ratio [95% CI] | p | Odds ratio [95% CI] | p | Odds ratio [95% CI] | p | Odds ratio [95% CI] | p | Odds ratio [95% CI] | p |
| **Sex** |  |  |  |  |  |  |  |  |  |  |
| Women | Ref |  | Ref |  | Ref |  | Ref |  | Ref |  |
| Men | **2.1 [1.9-2.3]** | **<0.001** | **2.1 [1.9-2.2]** | **<0.001** | **1.8 [1.7-2.0]** | **<0.001** | 1.0 [0.9-1.2] | 0.5 | **1.6 [1.5-1.8]** | **<0.001** |
| **Age** |  |  |  |  |  |  |  |  |  |  |
| 50-54 years | Ref |  | Ref |  | Ref |  | Ref |  | Ref |  |
| 55-59 years | **1.7 [1.5-1.9]** | **<0.001** | **1.6 [1.5-1.9]** | **<0.001** | **1.5 [1.3-1.7]** | **<0.001** | 1.0 [0.8-1.3] | 0.9 | 1.1 [1.0-1.3] | 0.08 |
| 60-64 years | **2.0 [1.8-2.3]** | **<0.001** | **2.0 [1.7-2.2]** | **<0.001** | **1.6 [1.4-1.8]** | **<0.001** | **1.3 [1.1-1.6]** | **0.007** | **1.3 [1.2-1.5]** | **<0.001** |
| 65-69 years | **2.3 [2.0-2.5]** | **<0.001** | **2.2 [2.0-2.5]** | **<0.001** | **1.7 [1.5-1.9]** | **<0.001** | 1.2 [1.0-1.5] | 0.06 | **1.2 [1.1-1.4]** | **0.004** |
| 70-74 years | **2.5 [2.2-2.8]** | **<0.001** | **2.4 [2.1-2.7]** | **<0.001** | **1.8 [1.6-2.1]** | **<0.001** | 1.0 [0.8-1.2] | 0.7 | 1.0 [0.9-1.2] | 0.6 |
| **Screening history** |  |  |  |  |  |  |  |  |  |  |
| No screening | Ref |  | Ref |  | Ref |  | Ref |  | Ref |  |
| Colonoscopy | **0.4 [0.3-0.5]** | **<0.001** | **0.4 [0.3-0.5]** | **<0.001** | **0.3 [0.2-0.3]** | **<0.001** | **0.6 [0.4-0.9]** | **0.03** | **0.6 [0.4-0.7]** | **<0.001** |
| gFOBT | 0.9 [0.9-1.0] | 0.2 | 0.9 [0.9-1.0] | 0.2 | **0.9 [0.8-1.0]** | **0.03** | 1.0 [0.8-1.1] | 0.7 | 0.9 [0.8-1.1] | 0.3 |
| FIT | **0.8 [0.7-0.9]** | **<0.001** | **0.8 [0.7-0.9]** | **<0.001** | **0.7 [0.6-0.7]** | **<0.001** | 1.0 [0.8-1.2] | 0.7 | 0.9 [0.8-1.1] | 0.4 |
| **Fecal**<br>**hemoglobin**<br>**concentration** |  |  |  |  |  |  |  |  |  |  |
| 10 µg/g | **1.02 [1.02-1.03]** | **<0.001** | **1.02 [1.01-1.03]** | **<0.001** | **1.06 [1.05-1.07]** | **<0.001** | 1.01 [1.00-1.02] | 0.3 | 1.00 [0.99-1.00] | 0.4 |
| **CIR** |  |  |  |  |  |  |  |  |  |  |
| ≥ 95% | Ref |  | Ref |  | Ref |  | Ref |  | Ref |  |
| 90 – 95% | 0.8 [0.6-1.1] | 0.2 | 0.8 [0.6-1.1] | 0.2 | 1.0 [0.7-1.3] | 0.7 | 0.7 [0.4-1.3] | 0.3 | 0.9 [0.6-1.4] | 0.6 |
| < 90 % | **0.5 [0.3-0.8]** | **0.004** | **0.5 [0.3-0.8]** | **0.004** | 0.7 [0.5-1.1] | 0.2 | 0.4 [0.1-1.1] | 0.07 | 0.8 [0.4-1.6] | 0.6 |

## DISCUSSION

### Main findings

The overall ADR in FIT-positive colonoscopies was 57.6% in this study, higher than ADRs reported in other FIT screening programs (43.5% to 51.5%),^15,16,26^ largely higher than those observed in gFOBT (35% to 47%)^26,27^ and colonoscopy screening programs (20% to 25%). ^7,28^ However, dramatic inter-endoscopist variation in adenoma detection and characterization were observed in our organized screening program supposed to offer to all invited persons an equally high-quality service. Overall, 35.9% of individuals were not being given the best possible chances as their colonoscopies were performed by 48.3% of endoscopists who had low CIRs and/or low ADRs. By contrast, 52.9% of individuals were being given the best possible chances as their colonoscopies were performed by 42.5% of endoscopists who were high detectors (NewADR ≥55%) maintaining good characterization ability (NNPDR ≤30%). We further found that a single indicator, NewADR, including proximal SLs and excluding CRCs, was enough to discriminate between high and low detectors, provided its target standard was raised to 55% to 70%. Last, we are the first to demonstrate that each 10 μg/g increase in fecal hemoglobin concentration was associated with a 2% increase in ADR.

### Alternative detection indicators

Overall, our correlations between neoplasia-related quality indicators were similar to the literature. There is no ideal detection indicator, the purpose of which is to assess and compare performances between individual endoscopists, but also between endoscopy centers and CRC screening strategies and programs. The most appropriate indicator is one which is actually measured in routine practice. It therefore has to be user-friendly for busy endoscopists, i.e. a unique global indicator that is easy to calculate and correlated with post-colonoscopy CRC risk and death. It should not be chosen on susceptibility to gaming but on reliable metrological properties. Especially for a worldwide indicator, as gaming is mostly linked with the payment system which differs between countries. All detection indicators are equally complicated to calculate, as they all depend on the pathology report, except for PDR and PR that can be automatically derived from administrative data. The correlations are strong between ADR and PDR or PR.^2,13^ Moreover, PR is significantly associated with risk of proximal post-colonoscopy CRC.^5^ However, the ratio to estimate ADR from PDR varies from 0.53 to 0.68 depending on studies and gender and varied from 0.61 to 0.93 in our study depending on the endoscopist.^13^ These large variations reflect the very different behaviors of endoscopists encountering a polyp, depending on many factors, such as education, training, experience, personality, time availability, withdrawal time, use of electronic chromoendoscopy and/or high-definition endoscopes, payment system, etc. Therefore, ADR cannot be estimated from PDR or PR using a unique conversion factor for the evaluation of an individual endoscopist. The conversion factor should be first evaluated individually by the endoscopist on a sample of 50 colonoscopies, and then used for the assessment of subsequent colonoscopies.^29^

Our results concerning SLs are similar to the literature. Our overall SSLDR was 3.9% (95%CI 3.6-4.3), significantly higher than reported in another large FIT screening program, estimated at 1.8% (95%CI 1.7-1.9).^30^ Likewise, our overall ProxSPDR was 7.6% (95%CI 7.2-8.1), within the range of 3% to 13% reported in screening colonoscopies.^11^ As others, we found a good correlation between SSLDR, ProxSLDR and ADR and all other detection indicators, broad inter-endoscopist variations (0% to 25%), and a significant ProxSLDR decrease in low-volume (<20 annual) and low-CIR (<90%) endoscopists.^11,30,31^ We further found a significant ProxSLDR decrease in individuals previously screened by colonoscopy (OR 0.6; 95%CI 0.4-0.9), whereas there was no significant difference in individuals previously screened by gFOBT and FIT.

### ADR benchmark

The ASGE/ACG Task Force on Quality in Endoscopy raised the recommended minimum target for ADR to 25% in 2015.^8^ This target was adopted by the ESGE in 2017^9^ and validated by three studies.^4,6,7^ For each 1% increase in ADR, a 3% reduction in post-colonoscopy CRC incidence and a 5% reduction in post-colonoscopy CRC mortality were observed.^6^ There is no evidence-based benchmark established for FOBT screening. A benchmark ADR of 35% is recommended for gFOBT screening in the English BCSP as in the previous French program.^10^ The US Multi-Society Task Force on CRC recommended a benchmark of 45% in men and 35% in women for FIT-positive colonoscopies (positivity threshold 20 μg/g).^20^ Two studies estimated that an ADR of 25% in screening colonoscopies in average risk individuals corresponded to ADRs of 49% and 55% in FIT-positive colonoscopies (positivity threshold 15 μg /g).^17,18^ The level of evidence is however low to moderate. Strong evidence could be derived from studies evaluating post-colonoscopy CRC risk and its association with ADR within CRC screening programs with FIT, but results will not be available for several years. Meanwhile, French recommendations adopted a benchmark of 45% based on our first FIT round. This low cut-off was chosen so that only 12.5% of endoscopists with very low ADRs – the greatest contributors to failure to prevent CRC – were considered low detectors. It is situated below our overall prevalence of adenomas (57.6%) and well below our true prevalence (70-80%) reached by very high detectors. We consider that a minimum of 45%, under which the risk of post-colonoscopy CRC would be prohibitive, should be a condition for acquiring and maintaining certification. However, we would recommend raising the minimum standard to 55%. The rationale is based on the dose-dependent approximately linear relationship between ADR and post-colonoscopy CRC risk, without a clear threshold.^6^ The more one detects adenomas, the less is the post-colonoscopy CRC risk. We chose 55% because all endoscopists having an ADR ≥55% had an MNA ≥0.8 (except one) (fig 1) and a ProxSLDR ≥4% (except three) (fig 2). Thus, a single indicator target, ADR ≥55%, would enable selection of high detectors for both conventional adenomas and proximal SLs, while avoiding the “one and done” pattern. Furthermore, the MNAPPC did not add any complementary information on colonoscopy quality.

For us, adopting the highest detectors’ performance level as the aspirational benchmark is not desirable because that level is achieved by detecting and removing all diminutive lesions, both adenomas and NNPs.^32,33^ ADR is the ideal tool to improve adenoma detection but should not lead to an endless race. There is no proof that the risk of post-colonoscopy CRC of very high detectors is significantly lower than that of high detectors. The overall rates of NNPs and diminutive polyps were much lower in our population-based study than in a single high level detector series (22.9% vs 35.7%, and 54.7% vs 75.0%, respectively, p<0.00001).^33^ “High-level detectors can produce a substantial economic burden of polyp resection and pathology charges for lesions with minimal clinical significance”.^33^ A maximum standard for the target ADR is thus desirable. To determine this maximum, we propose to adopt the true prevalence of “clinically relevant adenomas”, and not of “all adenomas”. We adopted 70% because NNPDR and NANSLDR were significantly higher in endoscopists having an ADR >70% than in those having an ADR ≤70%. Interestingly, using another method, Rex et al. proposed an aspirational target of 65-70% for FIT-positive colonoscopies, similar to our 55-70% proposition.^33^

We chose to present endoscopists’ individual ADRs using funnel plots as they avoid “the rather spurious ranking explicit in usual forest plots” (fig 4).^34^ Overall, 25/116 (21.6%) endoscopists were low detectors (<55%) and performed 2485 (18.5%) colonoscopies (of them 9/116 (7.8%) were very low detectors (<45%) and performed 664 colonoscopies (4.9%)), and 7/116 (6.0%) endoscopists were very high detectors (>70%) and performed 487 (3.6%) colonoscopies.

**Figure 4:**
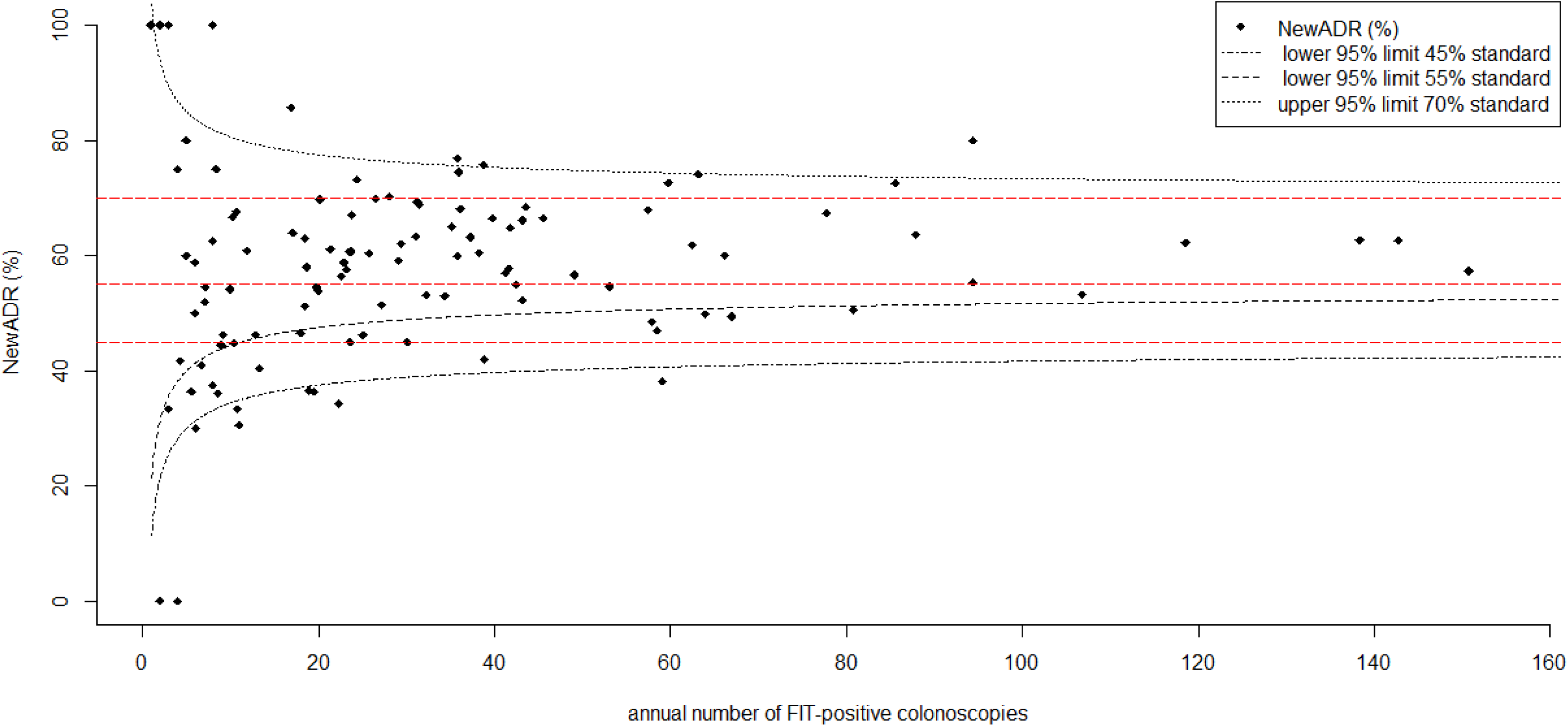
Funnel plots representing the adenoma detection rates of 116 endoscopists according to their annual fecal immunochemical test positive colonoscopy volume

### Method for calculating ADR

We did not count SSLs to calculate ADR.^8^ Authors rarely mention whether SSLs are included^5,35^ or excluded.^4,6,7,32,33,36^ The US recommendations exclude SSLs because most are not adenomas (no dysplasia).^8^ However, most are premalignant lesions (actually 9.4% of all premalignant lesions).^37^ Excluding them ignores completely the serrated pathway. One cannot simultaneously put forward arguments for promoting colonoscopy screening (e.g. against computed tomography colonography or FIT), i.e. emphasize the importance of detecting and removing proximal SLs, and promote a neoplasia-related quality indicator that ignores them. Another argument for excluding SSLs is that “pathologic distinction between SSP and HP is subject to marked interobserver variation in pathologic interpretation”.^8^ This has been true in the past, just as there has been very high inter-endoscopist variation in SL detection.^11^ However, there is no reason to believe pathologists could not make as much progress as endoscopists in recognizing SLs. Last, ADR (excluding SSLs) and NewADR (including proximal SLs instead of SSLs to avoid pathology interpretation difficulties) are equally difficult to calculate as they both rely on the pathology report. The difference between the two indicators was previously negligible, almost zero in the first gFOBT period, but became significant in the FIT period, and reached 8.7% in high SL detectors (fig 8). The difference is likely to grow with better detection of proximal SLs by endoscopists and their recognition by pathologists, so that proximal SLs cannot be ignored any more when calculating a single global detection indicator. We advocate the replacement of ADR by NewADR. As ADR has become a household name for gastroenterologists, we chose to preserve the term ADR when promoting NewADR, even if most proximal SLs are not adenomas.

We excluded colonoscopies displaying CRCs for the calculation of all indicators. Some authors count them when calculating ADR,^4,6,25,28^ most don’t. Interestingly, two^4,6^ of three^4-6^ landmark studies on association between ADR or PR and post-colonoscopy CRC risk counted CRCs. They should probably not be counted any more. As others, we found that the CRC detection rate is not endoscopist dependent. Whereas ADR wouldn’t change significantly if CRCs were included in screening colonoscopies displaying CRC in about 1% of cases, ADR would increase significantly in enriched FOBT-positive colonoscopies displaying CRC in >5% of cases. In our FIT period, the overall ADR would increase significantly by 2.3% if CRCs were included (from 0% to 4.9% depending on the endoscopist).

Another unanswered question is whether incomplete colonoscopies and inadequate bowel preparation colonoscopies have to be excluded or not for ADR calculation. US and European recommendations do not specify this point.^8,9^ Some studies exclude incomplete colonoscopies, ^5,13,17,19,25,32^ most don’t.^7,15,18,27,28,30,31,33,35^ We did not exclude them. We found no significant difference between the two calculation methods. Some studies exclude colonoscopies with poor bowel preparation,^4,7,17,19,25,36^ most don’t.^5,7,15,18,27,28,30,31,33,35^ We did not exclude them. It has been demonstrated that ADRs are significantly higher in colonoscopies with intermediate- and high-quality bowel preparation than in those with poor preparation.^38^

These observations indicate that there is an imperative and urgent need for a worldwide consensus on standardized indicators for the assessment of the benefit-risk balance of screening strategies and programs. An international task force should be created to formulate a series of recommendations, standardize definitions and categories and develop standardized methodology to calculate neoplasia-related and other quality indicators, for the same reasons and in the same way as were established a consensus on post-colonoscopy CRC and the CONSORT statement.^39^

### Colonoscopy volume

Two issues arise regarding the annual colonoscopy volume of endoscopists: is a minimum number of colonoscopies necessary 1) for adequate evaluation of ADR, and 2) to ensure high-quality performance?

1. The ESGE recommends continuous monitoring of ADR in all colonoscopies performed in patients aged 50 years or older.^9^ However, continuous monitoring is currently the exception because it remains a time-consuming process in the absence of modern endoscopy reporting systems permitting automatic entry of pathology results into databases. In eager expectation of the ideal computerized tool, approximate evaluation is better than no evaluation. An accurate measure would be mandatory if the result were used for applying reward or penalty. Such is not the case. Today, French endoscopists do not measure their ADR in all colonoscopies performed in patients aged 50 years or older. The only way a number of them are informed about their ADR is periodic feedback from some centers in charge of organizing the CRC screening program with FIT. In Alsace, ADRs and their 95%CIs are transmitted annually to all endoscopists. Since 2020, they also receive their ProxSLDRs and NNPDRs. Failing continuous monitoring, the ESGE guidelines recommend a yearly audit of a sample of 100 consecutive colonoscopies.^9^ However, waiting for 100 colonoscopies for a better accuracy, as reflected by a smaller 95%CI, would translate into prohibitive delays for a number of endoscopists.^40^ We propose that the minimum number of colonoscopies needed to estimate ADR should be reduced to 30, enabling early feedback for root cause analysis and subsequent implementation of improvement measures when necessary. In Alsace, only 4.3% of endoscopists perform ≥100 FIT-positive colonoscopies annually, the median time-frame for reaching 100 colonoscopies being 23 months. The minimum number of colonoscopies per endoscopist needed for inclusion in previous studies about ADR varied from 20 to 100.^31^ As in most studies, we chose 30 as this number is the minimum necessary to achieve an approximate normal distribution.^4,7,15^ It allowed us to evaluate 69.0% of endoscopists within a four-year period, the median time-frame for reaching 30 FIT-positive colonoscopies being 9 months. How endoscopists performing <30 FIT-positive colonoscopies should be evaluated remains to be stated. However, one could wonder whether low-volume endoscopists should participate in organized CRC screening programs.
2. All certified community gastroenterologists participate in the French CRC screening program. We did not observe, as in most studies, any correlation between ADR and colonoscopy volume.^15,31,41^ By contrast, low-volume endoscopists (<20 annual FIT-positive colonoscopies) had significantly lower CIRs and ProxSLDRs and higher NANSLDRs than high-volume endoscopists, so that their benefit/risk and cost-effectiveness ratios of polypectomy were significantly lower. The association between low volume and lesser care quality is well demonstrated for some other complex endoscopic procedures such as endoscopic retrograde cholangio-pancreatography.^42^ These results prove England and The Netherlands right as their CRC screening programs are based on endoscopists’ accreditation.

### Characterization indicators

The correlation between detection and characterization indicators was moderate (ADR) to strong (ProxSLDR) in our study. Previous small single-center studies observed strong correlations between ADR and NANSLDR or NNPDR.^32,33,39^ In other words, the more endoscopists detected adenomas and proximal SLs, the lower their PPV for NPs, i.e. the lesser their characterization competency and greater the number of NNPs they removed. There is neither an indicator nor a benchmark recommended to estimate the proportion of NNPs removed during colonoscopy. Such an indicator would represent the ability of the endoscopist to differentiate NPs that have to be removed from NNPs that should be left *in situ* and to estimate indirectly the cost-effectiveness of polypectomy. The lower the proportion of NNPs removed, the better cost-effectiveness is. NNPDR and NANSLDR are potential candidates. They vary largely depending on the endoscopist.^35,36,43^ Literature is scarce in this field. Regardless of the indicator, our proportion of NNPs was lower than previously reported. Our overall NNPDR was 22.9%, significantly lower than the 28-29% observed in a single-center study (p<0.001).^43^ Likewise, our overall NANSLDR was 6.4%, significantly lower than the 9-10% observed in two small single-center studies (p<0.001).^35,36^ Technological progresses, such as high definition, electronic chromoendoscopy (e.g. NBI) or artificial intelligence, should allow increased characterization ability. Some authors, using new technologies, are able to discriminate NPs from NNPs in >90% of cases.^22,44^ Such high accuracy is however controversial and, in any case, community endoscopists are far from these performances.^22,23^ Moreover, NNPDR and NANSLDR increased continuously over time in our study, probably the perverse effect of the adenoma detection race, endoscopists being encouraged to improve their ADRs and thus being prompted to detect and remove as many polyps as possible. Feedback and training are thus desirable to increase characterization performances and diminish unnecessary risks and costs. For routine use, NANSLDR being too restrictive, we would recommend measuring periodically NNPDR and propose a maximum standard of 30% corresponding to the third quartile in our study (and an aspirational maximum standard of 25% corresponding to the second quartile).

### Strengths and limitations of the study

This was a large population- and community-based study, which is its main strength. Other strengths include prospectively collected data and a high-quality database, as evidenced by our 5% rate of unknown polyp histology. Our study is, to our knowledge, the first to analyze the association between neoplasia-related indicators and fecal hemoglobin concentration and screening history. Our study is not without limitations. Some factors influencing neoplasia yield were not included in our analysis. For instance, certain patient-related factors such as body mass index, smoking habits or quality of bowel preparation, endoscopist-related factors such as withdrawal time and technique, practice duration, endoscope-related factors such as instrument generation, and endoscopy center-related factors such as the existence of screening-dedicated sessions^15^ were not examined. This should however not modify our findings because the principal demographic features predictive of neoplasia at colonoscopy are age and gender, which were analyzed, and to a lesser extent family history of colorectal neoplasia, which was excluded from our screening program. Last, our standards are specific for our population and FIT positivity threshold (30 μg/g). Moreover, our FIT-positive population was not screening naïve as a gFOBT CRC screening program had been running for 8 to 12 years before. Standards should be raised for lower positivity thresholds and in naïve populations.

### Future research

Future research should be directed at measuring the correlation between ADR and the risk of post-colonoscopy CRC in FIT screening programs. The low ADRs observed in a number of endoscopists is an issue in organized CRC screening programs supposed to offer to the screened population a better quality assurance than opportunistic screening. Reduction of the percentage of low detectors should be a priority for quality improvement programs as they are the main cause of post-colonoscopy CRCs. Several studies have demonstrated that it is possible to improve performance through well-designed training programs.^45^ Endoscopists whose ADRs are <55% should improve their detection ability. Those whose ADRs exceed 55% should improve their characterization ability to leave *in situ* NNPs and improve the benefit/risk and cost-effectiveness balances of their procedures. Another field of research is the evaluation of polypectomy competency. We proposed elsewhere a new indicator to evaluate the latter and demonstrated that detection and polypectomy competency were correlated.^46^ And finally, the endoscopist’s ADR should be considered for determining the post-polypectomy surveillance schedule in order to address the paradox that the shortest surveillance intervals are recommended by endoscopists with the highest ADRs, and inversely. Today, none of the existing recommendations take into account the endoscopist’s ADR for the determination of the post-polypectomy surveillance schedule. Yet, a small retrospective study found that the ADR of the endoscopist who performed the baseline colonoscopy was the only independent predictor of risk for advanced neoplasia at follow-up.^47^

### Conclusion

A single indicator, ADR, is enough to assess the neoplasia yield of colonoscopy. There is no need for alternative indicators, provided both of two conditions are met: 1) proximal SLs are counted for its calculation, and 2) the desirable target standard is raised to a higher level than previously recommended. Therefore, we propose to replace ADR by NewADR, including proximal SLs and excluding CRCs. In the French organized CRC screening program with FIT (30 μg/g positivity cut-off), the desirable target standard should be established between 55% and 70% (65-80% in men, 45-60% in women) to maximize the benefit / risk balance of the program. We further propose to adopt the NNPDR for evaluation of the endoscopist’s characterization ability, an indirect estimation of the cost-effectiveness of polypectomy, and to set its maximum standard at 30%. In any case, owing to the heterogeneous mixture of methods displayed in the literature, there is an imperative need for an internationally recognized standardized way to calculate neoplasia-related quality indicators.

## Data Availability

All deidentified participant data are available upon reasonable request from IG

**Abbreviations**
ADR: Adenoma detection rate
AUC: Area under curve
A10+DR: Adenoma ≥ 10 mm detection rate
ASGE: American Society for Gastrointestinal Endoscopy
CIR: Cecal intubation rate
CRC: Colorectal cancer
DistHPDR: Distal hyperplastic polyp detection rate
ESGE: European Society of Gastrointestinal Endoscopy
FIT: fecal immunochemical test
gFOBT: Guaiac based fecal occult blood test
MNA: Mean number of adenomas per colonoscopy
MNAPPC: Mean number of adenomas per positive colonoscopy
MNP: Mean number of polyps per colonoscopy
NANSLDR: Non-adenomatous non-serrated lesion detection rate
NewADR: New adenoma detection rate
NP: Neoplastic polyp
NNP: Non-neoplastic polyp
NNPDR: Non-neoplastic polyp detection rate
PAP: Proportion of adenomas among polyps
PDR: Polyp detection rate
PPV: Positive predictive value
PR: Polypectomy rate
ProxSPDR: Proximal serrated polyp detection rate
SD: Standard deviation
SL: Serrated lesion
SSA/P: Sessile serrated adenoma/polyp
SSA/PDR: Sessile serrated adenoma/polyp detection rate

---

**Suppl file 1:** Characteristics of all polyps removed within the colorectal cancer screening program with fecal immunochemical test

**Suppl file 2:** Correlation between adenoma and polyp detection rates

**Suppl file 3:** Detection and characterization indicators during the three periods of the colorectal cancer screening program with guaiac-based fecal occult blood test and fecal immunochemical test (endoscopists having performed ≥30 colonoscopies)

**Suppl file 4:** Changes in detection and characterization indicators between guaiac-based fecal occult blood test and fecal immunochemical test screening periods

**Suppl file 5:** Multivariate analyses of factors influencing detection and characterization indicators during the two screening periods

## Acknowledgments

The authors thank all the gastroenterologists who participated in the organized CRC screening program. They also thank the staff of screening centers and the participating general practitioners and pathologists for their contributions.

## Notes

**Funding** This study was performed as part of a quality assurance program within the French national CRC screening program without dedicated funding. The sources of funding of the national program include the French statutory health insurance scheme (Assurance Maladie), the French Ministry of Health and some regional Administrations. They had no role in study design, data collection, analysis, and interpretation, or writing the report.

**Competing interests** Pr Gabriel Rahmi reports personal fees from Medtronic, Fujifilm, and grants from Norgine. None declared for other authors

### Competing Interest Statement

Pr Gabriel Rahmi reports personal fees from Medtronic, Fujifilm, and grants from Norgine. None declared for other authors

### Funding Statement

This study was performed as part of a quality assurance program within the French national CRC screening program without dedicated funding. The sources of funding of the national program include the French statutory health insurance scheme (Assurance Maladie), the French Ministry of Health and some regional Administrations. They had no role in study design, data collection, analysis, and interpretation, or writing the report.

### Author Declarations

This study was exempt from institutional review board approval because individual participants were not approached, only routinely collected data were utilised and all data were anonymized for the purposes of quality improvement within the screening program. The study was performed in accordance with the declaration of Helsinki.

## REFERENCES

1 Rex DK, Boland CR, Dominitz JA, et al. Colorectal Cancer Screening: Recommendations for Physicians and Patients From the U.S. Multi-Society Task Force on Colorectal Cancer. Gastroenterology 2017;153:307–323.

2 Denis B, Sauleau EA, Gendre I, et al. Measurement of adenoma detection and characterization during colonoscopy in routine practice: an exploratory study. Gastrointest Endosc 2011;74:1325–1336.

3 Denis B, Sauleau EA, Gendre I, et al. The mean number of adenomas per procedure should become the gold standard to measure the neoplasia yield of colonoscopy: a population-based cohort study. Dig Liver Dis 2014;46:176–181.

4 Kaminski MF, Regula J, Kraszewska E, et al. Quality indicators for colonoscopy and the risk of interval cancer. N Engl J Med 2010;362:1795–1803.

5 Baxter NN, Sutradhar R, Forbes SS, et al. Analysis of administrative data finds endoscopist quality measures associated with postcolonoscopy colorectal cancer. Gastroenterology 2011;140:65–72.

6 Corley DA, Jensen CD, Marks AR, et al. Adenoma detection rate and risk of colorectal cancer and death. N Engl J Med 2014;370:1298–1306.

7 Kaminski MF, Wieszczy P, Rupinski M, et al. Increased Rate of Adenoma Detection Associates With Reduced Risk of Colorectal Cancer and Death. Gastroenterology 2017;153:98-105.

8 Rex DK, Schoenfeld PS, Cohen J, et al. Quality indicators for colonoscopy. Gastrointest Endosc 2015;81:31–53.

9 Kaminski MF, Thomas-Gibson S, Bugajski M, et al. Performance measures for lower gastrointestinal endoscopy: a European Society of Gastrointestinal Endoscopy (ESGE) Quality Improvement Initiative. Endoscopy 2017; 49:378–397.

10 Quality assurance guidelines for colonoscopy. NHS Bowel Cancer Screening Program Publication N°6. February 2011. Available at: https://assets.publishing.service.gov.uk/government/uploads/system/uploads/attachment_data/file/427591/nhsbcsp06.pdf. Accessed October 2019.

11 East JE, Atkin WS, Bateman AC, et al. British Society of Gastroenterology position statement on serrated polyps in the colon and rectum. Gut 2017;66:1181–1196.

12 Zhao S, Wang S, Pan P, et al. Magnitude, Risk Factors, and Factors Associated With Adenoma Miss Rate of Tandem Colonoscopy: A Systematic Review and Meta-analysis. Gastroenterology 2019;156:1661–1674.

13 Williams JE, Holub JL, Faigel DO. Polypectomy rate is a valid quality measure for colonoscopy: results from a national endoscopy database. Gastrointest Endosc 2012;75:576–582.

14 Marcondes FO, Dean KM, Schoen RE, et al. The impact of exclusion criteria on a physician’s adenoma detection rate. Gastrointest Endosc. 2015;82:668–675.

15 Zorzi M, Senore C, Da Re F, et al. Quality of colonoscopy in an organised colorectal cancer screening program with immunochemical fecal occult blood test: the EQuIPE study (Evaluating Quality Indicators of the Performance of Endoscopy). Gut 2015;64:1389–1396.

16 Jensen CD, Corley DA, Quinn VP, et al. Fecal Immunochemical Test Program Performance Over 4 Rounds of Annual Screening: A Retrospective Cohort Study. Ann Intern Med 2016;164:456–463.

17 Hilsden RJ, Bridges R, Dube C, et al. Defining Benchmarks for Adenoma Detection Rate and Adenomas Per Colonoscopy in Patients Undergoing Colonoscopy Due to a Positive Fecal Immunochemical Test. Am J Gastroenterol 2016;111:1743–1749.

18 Cubiella J, Castells A, Andreu M, et al. COLONPREV study investigators. Correlation between adenoma detection rate in colonoscopy- and fecal immunochemical testing-based colorectal cancer screening programs. United European Gastroenterol J 2017;5:255–260.

19 Wong JCT, Chiu HM, Kim HS, et al. Adenoma detection rates in colonoscopies for positive fecal immunochemical tests versus direct screening colonoscopies. Gastrointest Endosc 2019;89:607–613.

20 Robertson DJ, Lee JK, Boland CR, et al. Recommendations on Fecal Immunochemical Testing to Screen for Colorectal Neoplasia: A Consensus Statement by the US Multi-Society Task Force on Colorectal Cancer. Gastroenterology 2017;152:1217–1237.

21 Crockett SD, Nagtegaal ID. Terminology, Molecular Features, Epidemiology, and Management of Serrated Colorectal Neoplasia. Gastroenterology 2019;157:949–966.

22 Mason SE, Poynter L, Takats Z, Darzi A, Kinross JM. Optical Technologies for Endoscopic Real-Time Histologic Assessment of Colorectal Polyps: A Meta-Analysis. Am J Gastroenterol 2019;114:1219–1230.

23 Rees CJ, Rajasekhar PT, Wilson A, et al. Narrow band imaging optical diagnosis of small colorectal polyps in routine clinical practice: the Detect Inspect Characterise Resect and Discard 2 (DISCARD 2) study. Gut 2017;66:887–895.

24 Denis B, Ruetsch M, Strentz P, et al. Short-term outcomes of the first round of a pilot colorectal cancer screening program with guaiac based fecal occult blood test. Gut 2007;56:1579–1584.

25 Anderson JC, Butterly LF, Weiss JE, et al. Providing data for serrated polyp detection rate benchmarks: an analysis of the New Hampshire Colonoscopy Registry. Gastrointest Endosc 2017;85:1188–1194.

26 Clark G, Strachan JA, Carey FA, et al. Transition to quantitative faecal immunochemical testing from guaiac faecal occult blood testing in a fully rolled-out population-based national bowel screening programme [published online ahead of print, 2020 Mar 31]. Gut. 2020;gutjnl-2019-320297.

27 Lee TJ, Rutter MD, Blanks RG, et al. Colonoscopy quality measures: experience from the NHS Bowel Cancer Screening Programme. Gut 2012;61:1050–7.

28 Corley DA, Jensen CD, Marks AR, et al. Variation of adenoma prevalence by age, sex, race, and colon location in a large population: implications for screening and quality programs. Clin Gastroenterol Hepatol 2013;11:172–180.

29 Schramm C, Scheller I, Franklin J, et al. Predicting ADR from PDR and individual adenoma-to-polyp-detection-rate ratio for screening and surveillance colonoscopies: A new approach to quality assessment. United European Gastroenterol J 2017;5:742–749.

30 Zorzi M, Senore C, Da Re F, et al. Detection rate and predictive factors of sessile serrated polyps in an organised colorectal cancer screening programme with immunochemical faecal occult blood test: the EQuIPE study (Evaluating Quality Indicators of the Performance of Endoscopy). Gut. 2017;66:1233–1240.

31 Sarvepalli S, Garber A, Rothberg MB, et al. Association of Adenoma and Proximal Sessile Serrated Polyp Detection Rates With Endoscopist Characteristics. JAMA Surg 2019;154:627–635.

32 Hilsden RJ, Rose SM, Dube C, et al. Defining and Applying Locally Relevant Benchmarks for the Adenoma Detection Rate. Am J Gastroenterol 2019;114:1315–1321.

33 Rex DK, Sullivan AW, Perkins AJ, et al. Colorectal polyp prevalence and aspirational detection targets determined using high definition colonoscopy and a high level detector in 2017. Dig Liver Dis 2020;52:72–78.

34 Spiegelhalter DJ. Handling over-dispersion of performance indicators. Qual Saf Health Care 2005;14:347–351.

35 Atia MA, Patel NC, Ratuapli SK, et al. Nonneoplastic polypectomy during screening colonoscopy: the impact on polyp detection rate, adenoma detection rate, and overall cost. Gastrointest Endosc 2015;82:370–375.

36 Melson J, Berger D, Greenspan M, et al. Maintaining low non-neoplastic polypectomy rates in high-quality screening colonoscopy. Gastrointest Endosc 2017;85:581–587.

37 Meester RGS, van Herk MMAGC, Lansdorp-Vogelaar I, Ladabaum U. Prevalence and Clinical Features of Sessile Serrated Polyps: A Systematic Review. Gastroenterology 2020;159:105–118.

38 Clark BT, Rustagi T, Laine L. What level of bowel prep quality requires early repeat colonoscopy: systematic review and meta-analysis of the impact of preparation quality on adenoma detection rate. Am J Gastroenterol 2014;109:1714–1724.

39 Rutter MD, Beintaris I, Valori R, et al. World Endoscopy Organization Consensus Statements on Post-Colonoscopy and Post-Imaging Colorectal Cancer. Gastroenterology 2018;155:909–925.

40 Do A, Weinberg J, Kakkar A, et al. Reliability of adenoma detection rate is based on procedural volume. Gastrointest Endosc 2013;77:376–380.

41 Forbes N, Boyne DJ, Mazurek MS, et al. Association Between Endoscopist Annual Procedure Volume and Colonoscopy Quality: Systematic Review and Meta-analysis [published online ahead of print, 2020 Mar 30]. Clin Gastroenterol Hepatol 2020;S1542-3565(20)30423-7.

42 Keswani RN, Qumseya BJ, O’Dwyer LC, et al. Association Between Endoscopist and Center Endoscopic Retrograde Cholangiopancreatography Volume With Procedure Success and Adverse Outcomes: A Systematic Review and Meta-analysis. Clin Gastroenterol Hepatol 2017;15:1866–1875.

43 Chen SC, Rex DK. Variable detection of nonadenomatous polyps by individual endoscopists at colonoscopy and correlation with adenoma detection. J Clin Gastroenterol 2008;42:704–707.

44 Chen PJ, Lin MC, Lai MJ, et al. Accurate Classification of Diminutive Colorectal Polyps Using Computer-Aided Analysis. Gastroenterology 2018;154:568–575.

45 Kaminski MF, Anderson J, Valori R, et al. Leadership training to improve adenoma detection rate in screening colonoscopy: a randomised trial. Gut 2016;65:616–624.

46 Denis B, Gendre I, Perrin P, et al. Management of large polyps in a colorectal cancer screening program with fecal immunochemical test: a population- and community-based observational study. medRxiv 2020.05.15.20103135; doi: https://doi.org/10.1101/2020.05.15.20103135

47 Mangas-Sanjuan C, Zapater P, Cubiella J, et al. Importance of endoscopist quality metrics for findings at surveillance colonoscopy: The detection-surveillance paradox. United European Gastroenterol J 2018;6:622–629.

